# Ethnic and socioeconomic differences in SARS-CoV-2 infection: prospective cohort study using UK Biobank

**DOI:** 10.1101/2020.04.22.20075663

**Authors:** Claire L Niedzwiedz, Catherine A O’Donnell, Bhautesh Dinesh Jani, Evangelia Demou, Frederick K Ho, Carlos Celis-Morales, Barbara I Nicholl, Frances Mair, Paul Welsh, Naveed Sattar, Jill P Pell, S Vittal Katikireddi

## Abstract

**Background:** Understanding of the role of ethnicity and socioeconomic position in the risk of developing SARS-CoV-2 infection is limited. We investigated this in the UK Biobank study.

**Methods:** The UK Biobank study recruited 40-70 year olds in 2006-2010 from the general population, collecting information about self-defined ethnicity and socioeconomic variables (including area-level socioeconomic deprivation and educational attainment). SARS-CoV-2 test results from Public Health England were linked to baseline UK Biobank data. Poisson regression with robust standard errors was used to assess risk ratios (RRs) between the exposures and dichotomous variables for: being tested, having a positive test and testing positive in hospital. We also investigated whether ethnicity and socioeconomic position were associated with having a positive test amongst those tested. We adjusted for covariates including age, sex, social variables (including healthcare work and household size), behavioural risk factors and baseline health.

**Results:** Among 428,225 participants in England, 1,474 had been tested and 669 tested positive between 16 March and 13 April 2020. Black, south Asian and white Irish people were more likely to have confirmed infection (RR 4.01 (95%CI 2.92-5.12); RR 2.11 (95%CI 1.43-3.10); and RR 1.60 (95% CI 1.08-2.38) respectively) and were more likely to be hospital cases compared to the White British. While they were more likely to be tested, they were also more likely to test positive. Adjustment for baseline health and behavioural risk factors led to little change, with only modest attenuation when accounting for socioeconomic variables. Socioeconomic deprivation and having no qualifications were consistently associated with a higher risk of confirmed infection (RR 2.26 (95%CI 1.76-2.90); and RR 1.91 (95%CI 1.53-2.38) respectively).

**Conclusions:** Some minority ethnic groups have a higher risk of confirmed SARS-CoV-2 infection in the UK Biobank study which was not accounted for by differences in socioeconomic conditions, measured baseline health or behavioural risk factors. An urgent response to addressing these elevated risks is required.

## Background

The Severe Acute Respiratory Syndrome coronavirus-2 (SARS-CoV-2) and its resulting disease (COVID-19) is spreading rapidly worldwide.^1^ A better understanding of the predictors of developing infection is essential for health service planning (e.g. ensuring adequate facilities for those most at risk), targeting prevention efforts (e.g. targeted shielding or surveillance) and for informing future modelling efforts. Age, male sex and pre-existing medical conditions are established predictors of adverse COVID-19 outcomes, as is excess adiposity,^2^ but the role of social determinants is poorly understood.^3,4^

Ethnicity and socioeconomic position strongly influence health outcomes for both infectious and non-communicable diseases. Previous pandemics have often disproportionately impacted ethnic minorities and socioeconomically disadvantaged populations.^5,6^ Early evidence suggests that the same may be occurring in the current SARS-CoV-2 pandemic but empirical research remains highly limited.^7^ It is highly plausible that infection risk will vary across these social groups. For example, socioeconomic disadvantage is linked to living in overcrowded housing and some ethnic groups are more likely to live in larger households^8^ – both of which potentially predispose to increased risk of infection, and to greater viral load.

Establishing the risk of developing infection across different social groups is challenging. A major issue is that information about ethnicity and socioeconomic position are often not well collected within routine health data. Furthermore, the size of the different social groups in the general population is also often not accurately known. The ideal approach to estimating infection risk across different social groups is to analyse data from a cohort study, but most existing cohort studies which include detailed information about ethnicity and socioeconomic position are subject to long delays in data being available for analysis and are too small to provide useful estimates of infection risk.

The UK Biobank study has carried out data linkage between its study participants and SARS-CoV-2 test results held by Public Health England. We therefore aimed to investigate the relationship between ethnicity, socioeconomic position and the risk of having confirmed SARS-CoV-2 infection in the population-based UK Biobank study.

## Methods

### Study design and participants

Data were obtained from UK Biobank (https://www.ukbiobank.ac.uk/), with the methods described in detail previously.^9^ In brief, over 502 000 community-dwelling individuals aged 37 to 73 years were recruited to the study during 2006 to 2010. Participants attended one of 22 assessment centres across England, Scotland and Wales. Data were collected on a range of topics including social and demographic factors, health and behavioural risk factors, using standardised questionnaires administered by trained interviewers and self-completion by computer.

Results of COVID-19 tests for UK Biobank participants, including confirmed cases, were provided by the Public Health England (PHE) microbiology database Second Generation Surveillance System and linked to UK Biobank baseline data.^10^ Data provided by PHE included the specimen date, specimen type (e.g. upper respiratory tract), laboratory, origin (whether there was evidence from microbiological record that the participant was an inpatient or not) and result (positive or negative). Data were available for the period 16 March 2020 to 14 April 2020.

Since data on test results were only available for England, we restricted the study population to people who attended UK Biobank baseline assessment centres in England. Participants who were identified as having died prior to 14 February 2018 from the linked mortality records provided by the NHS Information Centre and those who requested to withdraw from the study (N=26) were also excluded from the analysis. In addition to the analyses of the overall population, we also investigated positive test results among those who had been tested only. This allowed us to investigate the potential for bias due to differential testing between ethnic and socioeconomic groups. UK Biobank received ethical approval from the NHS National Research Ethics Service North West (11/NW/0382).

### Assessment of ethnicity and socioeconomic position

All exposures were derived from the baseline assessment centre data collection. Ethnicity was self-reported based on pre-defined categories into: white British, white Irish, other white background, south Asian, black (Caribbean or African), Chinese, mixed or other. Due to small numbers, analyses of the Mixed and Chinese groups were limited. In line with previous research, we also do not report results for the other group due to problems with interpretation of this highly heterogenous group.^11^

Socioeconomic position was assessed using two different measures recorded at the baseline visit. Area-level socioeconomic deprivation was assessed by the Townsend index (including measures of unemployment, non-car ownership, non-home ownership and household overcrowding) corresponding to the output area in which the respondent’s home postcode was recorded.^12^ Quartiles were derived from the index, where the lowest quartile represents the most advantaged and the highest the least advantaged. Highest education level usually remains stable throughout the adult life course and was assessed as 1) university or college degree, 2) A-levels or equivalent, 3) O-levels, General Certificate of Secondary Education (GCSE), vocational Certificate of Secondary Education (CSE) or equivalent, 4) other (e.g. National Vocational Qualifications or other professional qualifications), or 5) none of the above.^13^

### Ascertainment of SARS-CoV-2 outcomes

We defined our primary outcome as having a positive test within the Public Health England database available through linkage.^10^ This reflects confirmed infection but does not include symptomatic individuals who have not presented to the health service or not been tested, or asymptomatic cases. Some systemic differences exist in testing threshold. For example, healthcare workers may be more likely to be tested and therefore observed differences may reflect differences in testing practices. To investigate whether differential ascertainment was biasing our results, we studied three further outcomes. We identified positive cases that had their test taken while attending hospital (i.e. either Emergency Departments or as inpatients – hereafter referred to as hospital cases). This group is likely to reflect more severe illness and therefore is less likely to be subject to ascertainment bias. In addition, we investigated outcomes related to testing practice by assessing the risk of being tested in the overall population and testing positive amongst only those who had been tested. Higher levels of confirmed SARS-CoV-2 infection could arise from higher rates of testing amongst some population subgroups. However, if this were to occur, the likelihood of having a positive test would be lower amongst groups experiencing high rates of testing.

### Potential confounders and mediators

Age group (5-year age bands), sex and assessment centre were included as potential confounder variables in all statistical models. Country of birth (UK and Ireland) versus elsewhere was also included, given its influence on cultural practices.^14^ We also included several variables which could reflect potential confounding or mediation. Participants were asked about the title of their current or most recent job at baseline and these were converted to the Standard Occupational Classification (SOC 2000) by UK Biobank. Healthcare (and related) workers were identified from the SOC 2000 codes 22 (Health Professionals), 32 (Health and Social Welfare Associate Professionals), 118 (Health and Social Services Managers), 611 (Healthcare and Related Personal Services) and 9221 (Hospital porters).

Baseline health status was assessed using self-reported long-standing illness, health, disability or infirmity (yes or no) and the number of chronic health conditions self-reported from a pre-defined list of 43 conditions and top-coded at 4 or more, based on a previously published approach.^15^ Lifestyle factors included smoking (never, previous, current); body mass index (BMI) (weight/height^2^ derived from physical measurements and classified into underweight, normal weight, overweight, obese); and alcohol consumption (categorised into daily or almost daily, 3-4 times a week, once or twice a week, 1-3 times per month, special occasions, former drinker or never).

Other social variables were also considered. Employment status distinguished those in paid employment or self-employment, retired, looking after home and/or family, unable to work because of sickness or disability, unemployment or other. For those in work, manual versus non-manual occupation was assessed by asking participants to report whether their job involved heavy manual or physical work (never/rarely/sometimes versus usually/always). Housing tenure was categorised into owner-occupier or renter/other (including those who live in accommodation rent free, in a care home or sheltered accommodation). Urban/rural status was derived from data on the home area population density; UK Biobank combined each participant’s home postcode with data generated from the 2001 census from the Office of National Statistics. The number of people within a household was categorised into three groups: single person, two people and three or more people (which included those living in institutions, such as care homes).

### Statistical analyses

The association between the exposures (ethnicity and socioeconomic position) and the outcomes of interest (confirmed infection, hospital case, being tested and having a positive test amongst those tested) were explored using Poisson regression. Poisson regression was preferred over logistic regression to allow relative risks to be presented, rather than odds ratios which are often misinterpreted.^16^ Robust standard errors were used to ensure accurate estimation of 95% confidence intervals and p values. Missing data were excluded from the analysis. Sensitivity analyses were conducted running the key models for those with complete data on all variables. Statistical analysis was conducted using Stata/MP 15.1

To investigate ethnicity, we initially adjusted for age, sex and assessment centre (model 1) and then added country of birth (model 2). Subsequent models additionally adjusted for variables which we hypothesised were likely to be at least partially mediating rather than confounding variables. Model 3 adjusted for model 2 variables and for being a healthcare worker. Model 4 additionally adjusted for social variables (namely urbanicity, number of people per household, highest education level, deprivation, tenure status, employment status, manual work); model 5 was adjusted for model 2 plus health status variables (self-rated health, number of chronic conditions and limiting longstanding illness or disability); model 6 was adjusted for model 2 plus behavioural risk factors (smoking, alcohol consumption and BMI); and model 7 was adjusted for all aforementioned covariates.

We followed a similar approach to explore the role of deprivation and education level. Model 1 was adjusted for age, sex and assessment centre; model 2 added ethnicity and country of birth; model 3 also adjusted for the social variables (as above); model 4 adjusted for model 2 plus health status variables; model 5 was adjusted for model 2 plus behavioural risk factors; and model 6 was adjusted for all previous covariates.

## Results

Most of the baseline UK Biobank sample in England was white British, with the next largest groups being other white, white Irish and then south Asian and black (Table 1). Approximately one-third (32.6%) of the sample had a degree and 16.5% had no formal qualifications.

**Table 1.** Description of the study population

| **Age group** | **N** | **%** |
| --- | --- | --- |
| 37-44 | 45,633 | 10.66 |
| 45-49 | 57,353 | 13.39 |
| 50-54 | 65,746 | 15.35 |
| 55-59 | 77,370 | 18.07 |
| 60-64 | 103,493 | 24.17 |
| 65-69 | 76,634 | 17.90 |
| 70+ | 1,996 | 0.47 |
| **Sex** | **N** | **%** |
| Female | 235,149 | 54.91 |
| Male | 193,076 | 45.09 |
| **Ethnicity** | **N** | **%** |
| White British | 375,366 | 88.16 |
| White Irish | 10,822 | 2.54 |
| White Other | 14,365 | 3.37 |
| Mixed | 2,661 | 0.62 |
| South Asian | 9,246 | 2.17 |
| Black | 7,726 | 1.81 |
| Chinese | 1,402 | 0.33 |
| Other | 4,204 | 0.99 |
| **Country of birth** | **N** | **%** |
| UK & Ireland | 390,262 | 91.48 |
| Elsewhere | 36,358 | 8.52 |
| **Number in household** | **N** | **%** |
| 1 | 77,080 | 18.12 |
| 2 | 198,098 | 46.57 |
| 3+ | 150,239 | 35.32 |
| **Education Level** | **N** | **%** |
| College or University degree | 136,866 | 32.64 |
| A levels/AS levels or equivalent | 47,381 | 11.30 |
| O levels/GCSEs/CSEs or equivalent | 115,789 | 27.62 |
| Other | 49,689 | 11.85 |
| None of the above | 69,552 | 16.59 |
| **Deprivation (quartile)** | **N** | **%** |
| 1 | 107,625 | 25.16 |
| 2 | 107,394 | 25.11 |
| 3 | 107,022 | 25.02 |
| 4 | 105,683 | 24.71 |
| **Housing tenure** | **N** | **%** |
| Owner | 377,394 | 88.96 |
| Rent/Other | 46,847 | 11.04 |
| **Urban/Rural** | **N** | **%** |
| Urban | 363,563 | 85.72 |
| Rural | 60,569 | 14.28 |
| **Employment status** | **N** | **%** |
| In paid employment or self-employed | 248,602 | 58.41 |
| Retired | 138,955 | 32.65 |
| Looking after home and/or family | 12,131 | 2.85 |
| Unable to work because of sickness or d | 12,870 | 3.02 |
| Unemployed | 7,367 | 1.73 |
| Other | 5,675 | 1.33 |
| **Manual occupation** | **N** | **%** |
| Non-manual | 214,443 | 50.42 |
| Manual | 33,850 | 7.96 |
| Not in employment | 176,998 | 41.62 |
| **Health care worker** | **N** | **%** |
| No | 221,679 | 52.11 |
| Yes | 26,713 | 6.28 |
| Not in employment | 176,998 | 41.61 |
| **Long-standing illness, disability or infirmity** | **N** | **%** |
| No | 284,706 | 68.31 |
| Yes | 132,078 | 31.69 |
| **Number of long-term conditions** | **N** | **%** |
| 0 | 159,476 | 37.46 |
| 1 | 141,121 | 33.15 |
| 2 | 75,814 | 17.81 |
| 3 | 31,808 | 7.47 |
| 4+ | 17,536 | 4.12 |
| **Self-reported health** | **N** | **%** |
| Excellent | 69,282 | 16.29 |
| Good | 249,382 | 58.64 |
| Fair | 89,048 | 20.94 |
| Poor | 17,580 | 4.13 |
| **Body Mass Index (BMI)** | **N** | **%** |
| Underweight (<18.5) | 2,145 | 0.50 |
| Normal weight (18.5-24.9) | 140,102 | 32.91 |
| Overweight (25.0-29.9) | 181,011 | 42.52 |
| Obese (>=30.0) | 102,430 | 24.06 |
| **Smoking status** | **N** | **%** |
| Never | 235,711 | 55.37 |
| Previous | 147,235 | 34.59 |
| Current | 42,745 | 10.04 |
| **Alcohol consumption** | **N** | **%** |
| Daily or almost daily | 87,430 | 20.48 |
| Three or four times a week | 99,058 | 23.20 |
| Once or twice a week | 109,562 | 25.66 |
| One to three times a month | 47,720 | 11.18 |
| Special occasions only | 49,165 | 11.52 |
| Former drinker | 14,842 | 3.48 |
| Never | 19,144 | 4.48 |

Data for 1474 participants who had been tested for SARS-CoV-2 between 16 March and 13 April 2020 were linked to UK Biobank baseline data and available for analysis (N=428,225) (Additonal file Figure S1 for flowchart). Of these, 669 participants received a positive test result from a total of 2724 tests. 572 received a positive test while attending hospital, suggesting more severe illness. The geometric mean number of tests performed per participant tested was 1.63 (95% CI: 1.59 to 1.67), with relatively small differences in the numbers of tests by ethnicity and socioeconomic position.

In comparison to the white British majority ethnic group, several ethnic minority groups had a higher risk of testing positive for SARS-CoV-2 infection and also testing positive while attending hospital (Figure 1 and Additonal file: Table S3a). Black participants had the highest risk (RR 4.01 (95%CI 2.92-5.52)), with adjustment for country of birth resulting in little attenuation (RR 3.70 (95%CI 2.50-5.48)); adjustment for a history of being a healthcare worker (RR 3.35 (95%CI 2.24-5.00)) and for social factors (including measures of socioeconomic position) did additionally attenuate the risk (RR 2.45 (95%CI 1.57-3.81)). South Asians also had an elevated risk (RR 2.11 (95%CI 1.43-3.10) in model 1), with a similar pattern of attenuation as for the black ethnic group. In contrast, the white Irish group had a consistent modestly elevated risk of having a positive test (RR 1.60 (95%CI 1.08-2.38)) which did not change with adjustment for covariates. The Chinese group had imprecisely estimated risk ratios due to smaller numbers. The pattern of findings for hospital cases was similar, suggesting that the higher testing rates amongst certain ethnic groups in the community were not skewing the results. Similarly, analyses of the likelihood of testing positive amongst those who had been tested was often higher or the same in these ethnic groups (Table 2), whereas a lower risk would have suggested differentially high testing.

**Table 2.**
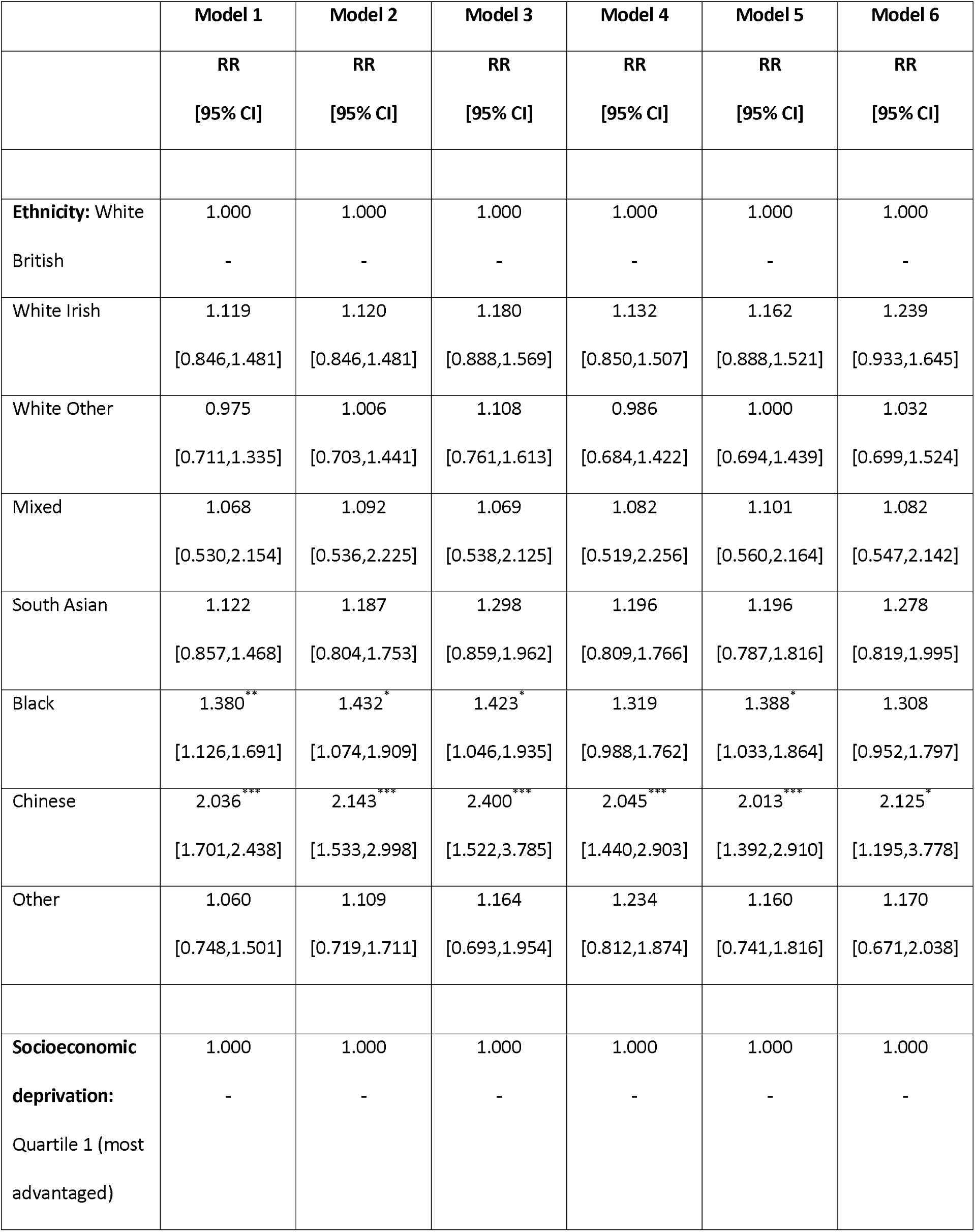
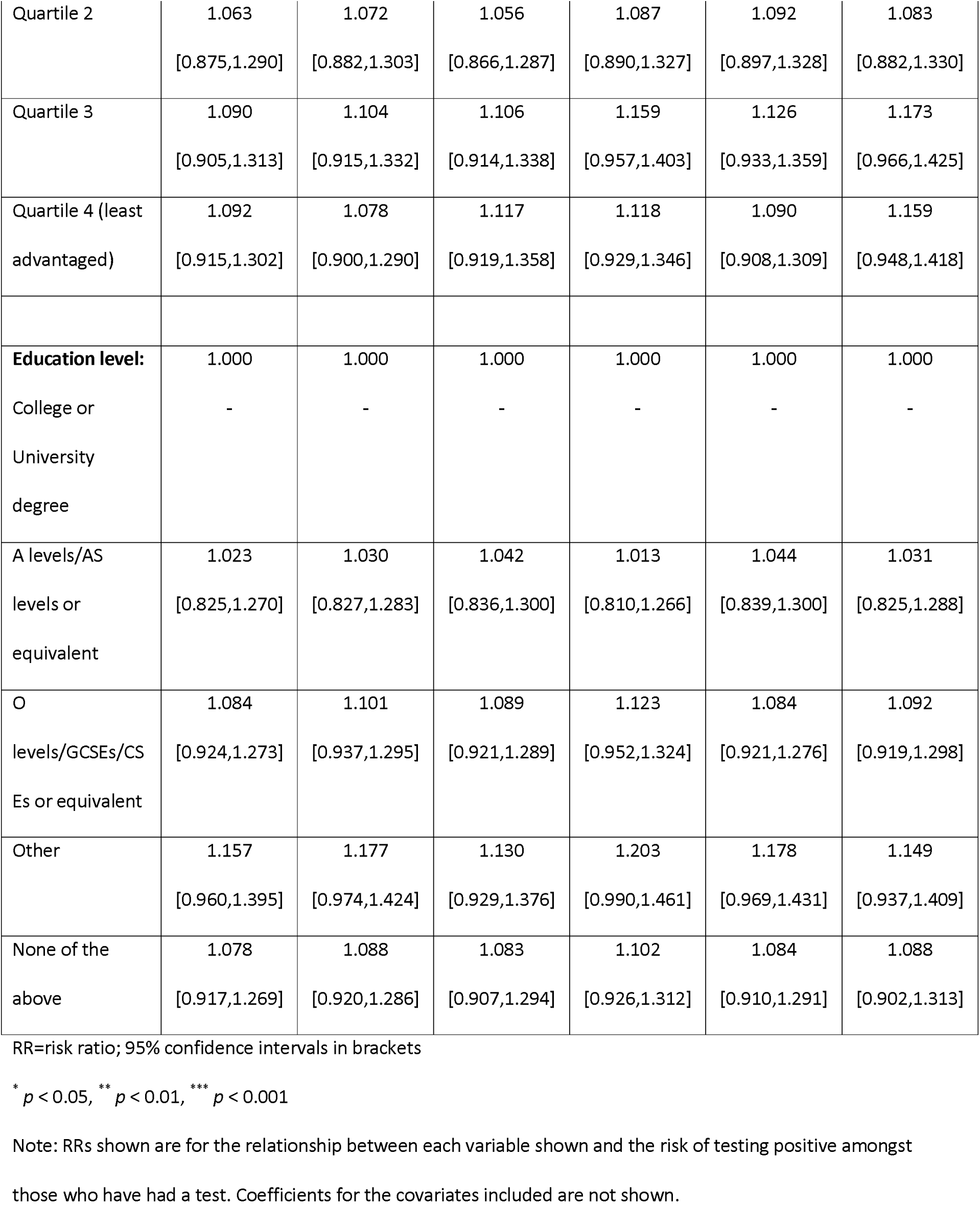
Risk ratios for testing positive for SARS-CoV-2 amongst those tested (N=1,474) in UK Biobank Model 1: Adjusted for age, sex, assessment centre Model 2: 1 + ethnicity, country of birth Model 3: 2 + education level, household size, socioeconomic deprivation, housing tenure, urbanicity, employment status, manual occupation, healthcare worker Model 4: 2 + limiting illness/disability, number of chronic conditions, self-rated health Model 5: 2 + Body Mass Index, smoking status, alcohol consumption Model 6: All of the above covariates

**Figure 1:**
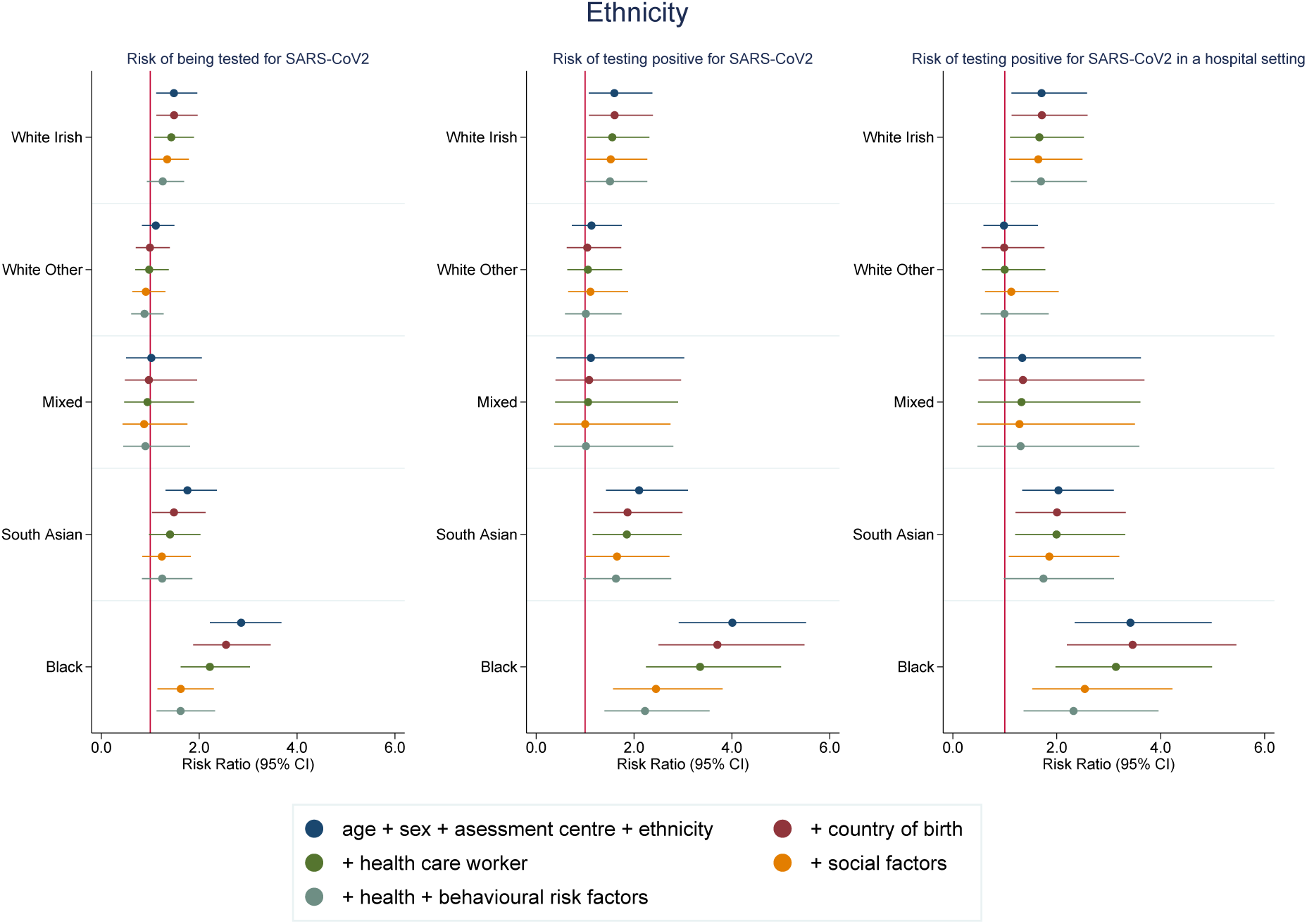
Risk ratios for associations between ethnicity (White British as reference category) and SARS-CoV-2. Model 1: Age, sex and assessment centre Model 2: Model 1 + country of birth Model 3: Model 2 + healthcare worker Model 4: Model 3 + social variables (urbanicity, number of people per household, highest education level, deprivation, tenure status, employment status, manual work) Model 5: Model 4 + health status variables (self-rated health, number of chronic conditions and limiting longstanding illness) + behavioural risk factors (smoking, alcohol consumption and BMI)

In comparison to the most socioeconomically advantaged quartile, living in a disadvantaged area (according to the Townsend deprivation score) was associated with a higher risk of confirmed infection, particularly for the most disadvantaged quartile (RR 2.48 (95%CI 1.95-3.16)) (Figure 2 and Additonal file: Table S4a). Differences in ethnicity and country of birth, social factors, baseline health and behavioural risk factors all moderately attenuated the association in the most disadvantaged quartile. Socioeconomic deprivation was also associated with hospital cases. While testing was again more likely, the risk of being diagnosed positive amongst those tested also tended to be higher, rather than lower (Table 2).

**Figure 2:**
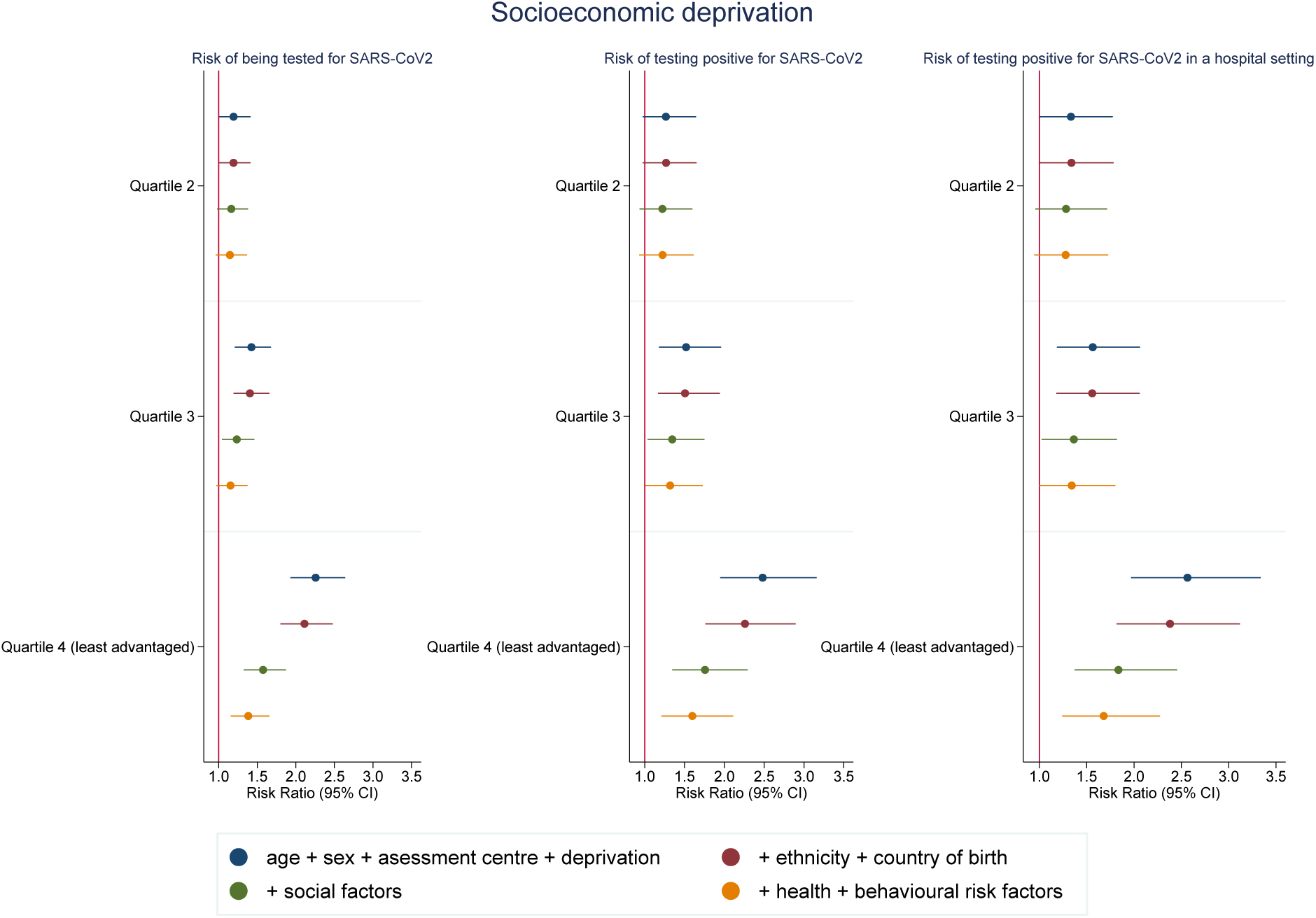
Risk ratios associations between Townsend deprivation score quartile (most advantaged as reference category) and SARS-CoV-2. Model 1: Age, sex and assessment centre Model 2: Model 1 + ethnicity + country of birth Model 3: Model 2 + social variables (healthcare worker, urbanicity, number of people per household, highest education level, tenure status, employment status, manual work) Model 4: Model 3 + health status variables (self-rated health, number of chronic conditions and limiting longstanding illness) + behavioural risk factors (smoking, alcohol consumption and BMI)

Analyses by education also showed a higher risk of confirmed SARS-CoV-2 infection with lower levels of education (RR 1.95 (95%CI 1.56-2.43) for no qualifications compared to degree level educated) (Figure 3 and Additional file: Table S5a). While adjustment for ethnicity and country of birth made little difference to the association, adjustment for social factors, baseline health and behavioural risk factors all attenuated the association somewhat (RR 1.41 (95%CI 1.09-1.82) in fully adjusted model). We again observed a similar pattern in hospital cases and found little evidence of increased testing amongst the less educated groups (Figure 3 and Table 2). We repeated analyses for those who had complete data on all variables (N= 392,116), but this made little difference to the substantive results (Additional file 1: Figures S2-S4).

**Figure 3:**
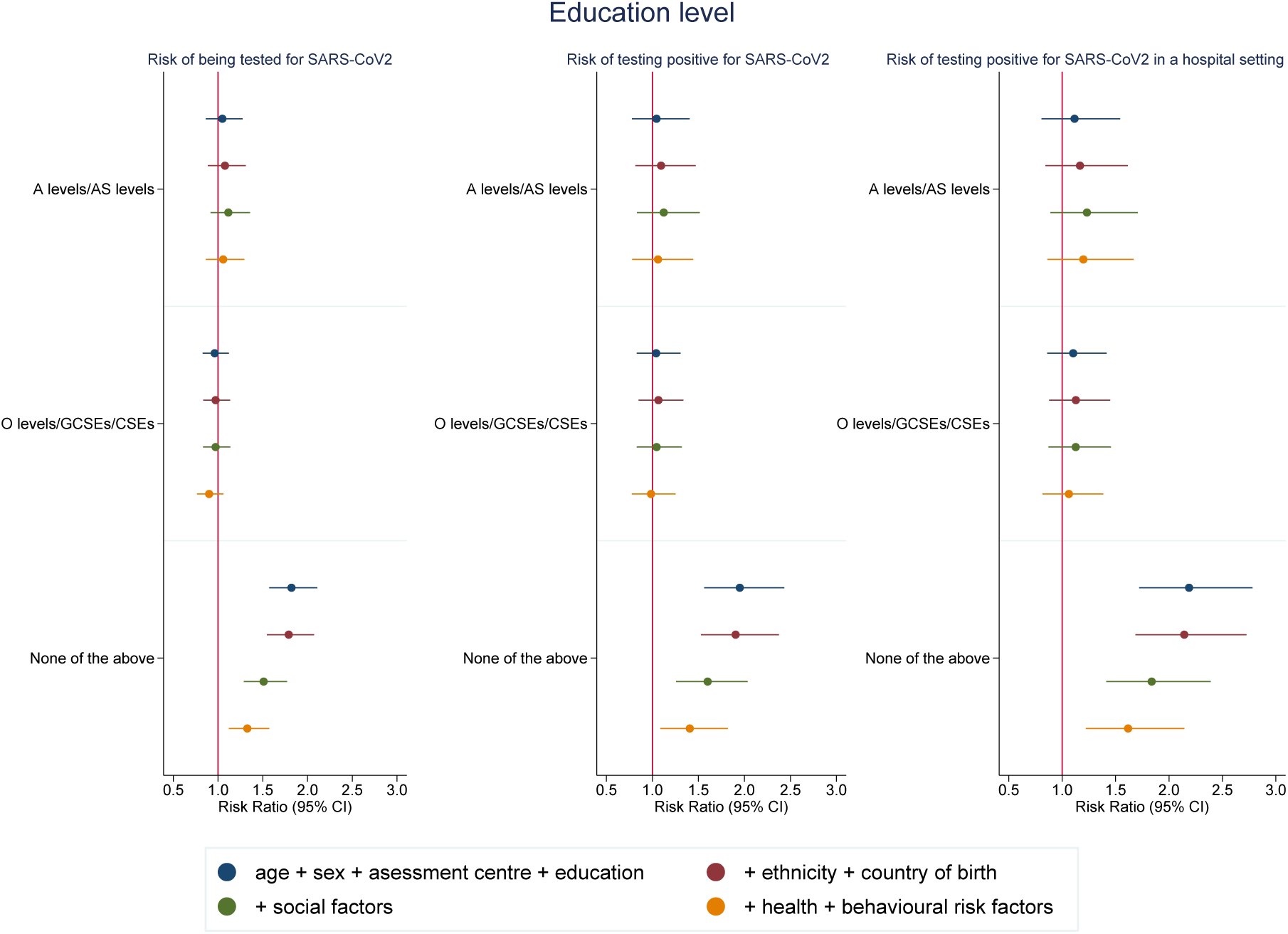
Risk ratios for associations between highest educational level (degree educated as reference category) and SARS-CoV-2. Model 1: Age, sex and assessment centre Model 2: Model 1 + ethnicity + country of birth Model 3: Model 2 + social variables (healthcare worker, urbanicity, number of people per household, deprivation, tenure status, employment status, manual work) Model 4: Model 3 + health status variables (self-rated health, number of chronic conditions and limiting longstanding illness) + behavioural risk factors (smoking, alcohol consumption and BMI)

## Discussion

Several ethnic minority groups had a higher risk of both being diagnosed and testing positive in a hospital setting with laboratory-confirmed SARS-CoV-2 infection in the UK Biobank study. The black, south Asian and white Irish ethnic groups were found to be at greatest risk. Similarly, measures of socioeconomic disadvantage (area-based deprivation and lower education) were also associated with an increased risk of having confirmed infection and being a hospital case. For both ethnicity and socioeconomic position, we did not find evidence that these patterns were likely to be due to differential ascertainment, since although the likelihood of testing was increased, the likelihood of a positive test was, if anything, higher among ethnic minorities who had been tested. Ethnic differences in infection risk did not appear to be fully accounted for by differences in pre-existing health, behavioural risk factors or country of birth measured at baseline. Furthermore, socioeconomic differences appeared to make a modest contribution to these ethnic differences.

Our study has several important strengths. First, by using a well characterised cohort study, we can identify a clearly defined population at risk of experiencing SARS-CoV-2 infection. By combining data linkage with a large sample size, this has allowed us to provide empirical data from this pandemic in a timely fashion. Ethnicity was collected using self-report which is widely considered to be a gold-standard approach^17^ and the availability of a large dataset has allowed us to provide empirical data on this crucial policy priority in a timely fashion, including a more nuanced appreciation of the risks of infection within different members of the white majority population, as well as minority ethnic populations. Our investigation of socioeconomic position has similarly benefited from being able to study different measures and assess the pattern of findings across these. The detailed data collected in this cohort has also allowed us to investigate the extent to which observed inequalities are potentially mediated by a wide range of factors, including behavioural risk factors, pre-existing health status and other social variables.

However, several potential limitations should be noted. Ascertainment bias is potentially problematic and could arise in several ways, including differential healthcare seeking, differential testing and differential prognosis. Even so, we have been unable to find any evidence to suggest that differential healthcare seeking or testing would explain the observed pattern of findings. Increased ascertainment amongst ethnic minorities would be expected to result in a lower proportion of confirmed cases amongst those tested whereas we observed the opposite. One possibility that remains is that some ethnic and socioeconomic groups have a poorer prognosis and are therefore more likely to be admitted to hospital and therefore to be tested. However, if this were the case, the issue of more adverse outcomes among these groups remains concerning. Other limitations include the non-representativeness of the UK Biobank study population, with those who were more advantaged being more likely to participate and ethnic minorities less well represented. There is therefore the potential that the findings in our study may not reflect the broader UK population.^18^ However, empirical research has found that this does not result in substantial bias in measures of association in the UK Biobank study.^19^ We have also been unable to fully exclude all deaths that occurred prior to the pandemic, due to lack of up-to-date linkage to mortality records at present. Our exposure data were collected some years ago and it is therefore likely that pre-existing health, risk factors and some social variables have changed, although generally most risk factors track throughout life. Being a healthcare worker was also ascertained at baseline, although many who stopped employment in this area have now returned to work. Lastly, we have not explored the role of specific health conditions such as asthma, diabetes and high blood pressure, which have been shown to be associated with a higher risk of severe outcomes^3,20^ and are more prevalent amongst socioeconomically disadvantaged groups and some ethnic minority groups.^21,22^ However, these are likely to operate as mediators rather than confounders.

Administrative data from health services has recently suggested an increased risk of severe COVID-19 disease within ethnic minority groups. The UK’s Intensive Care National Audit & Research Centre (ICNARC) analysed data on 5,578 patients admitted to critical care up to 16^th^ April 2020 and found black and Asian people comprised a high proportion of total patients (11.2% and 14.9% respectively), although it was unclear whether these higher percentages were biased by most cases being initially seen in areas with high BME proportions.^23^ Similarly, data from the US Centers for Disease Control and Prevention also suggest a higher risk amongst Black or African American people, but information on race was missing for approximately two-thirds of those diagnosed.^24^ Academic research on this topic has been limited to date. An ecological study of US counties has suggested that more socially vulnerable areas (which included greater numbers of people with socioeconomic disadvantage and ethnic minorities) were associated with higher COVID-19 case fatality rates.^25^ Our study adds substantially to the evidence by finding that ethnicity appears to be an important predictor of laboratory-confirmed SARS-CoV-2 infection that is only partly attenuated by a large range of potential mediators (such as socioeconomic position), as well as addressing concerns about numerator-denominator bias.

Our results suggest there is an urgent need for further research on how SARS-CoV-2 infection affects different ethnic and socioeconomic groups. Our findings warrant replication in other datasets, ideally including representative samples and across different countries. As the pandemic evolves, there is a need to monitor infection and disease outcomes by ethnicity and socioeconomic position. However, data to allow this disaggregation is often not available – record linkage could potentially help address this gap, particularly in settings where administrative register data are available. Given the differences in health risks across occupational groups^26^, understanding the risks that the full range of key workers experience is also required. Lastly, other social groups, such as homeless people, prisoners and undocumented migrants, experience severe disadvantage and research is necessary to study these highly vulnerable populations too.^27,28^

## Conclusions

The limited evidence available suggests that some ethnic minority groups, particularly black and south Asian people, are particularly vulnerable to SARS-CoV-2 infection. Socioeconomic disadvantage and poorer pre-existing health do not explain all this elevated risk. There is therefore a need to determine exactly why this increased risk occurs. An immediate policy response is required to ensure the health system is responsive to the needs of ethnic minority groups. This should include ensuring that health and care workforces, which often rely on workers from minority ethnic populations, have access to the necessary protective personal equipment (PPE) to ensure they can work safely. Timely communication of guidelines to reduce the risk of being exposed to the virus is also required in a range of languages.^29^ Previous evidence suggests ethnic minorities in the UK tend to receive reasonably equitable care in many, but not all, areas.^30^ However, this is not the case in many other countries (such as the US) where the adverse consequences of SARS-CoV-2 infection may be even worse. SARS-CoV-2 therefore has the potential to substantially exacerbate ethnic and socioeconomic inequalities in health^31^, unless steps are taken to mitigate these inequalities. The data from this study may be helpful to inform allocation of more aggressive therapies in people with severe disease, or targeting preventative vaccination to at risk groups, once evidence for such approaches becomes available.

## Data Availability

Data are available from UK Biobank.

https://www.ukbiobank.ac.uk/

## List of abbreviations

BMI: Body Mass Index
CI: Confidence interval
COVID-19: Coronavirus-19
CSE: Certificate of Secondary Education
GCSE: General Certificate of Secondary Education
ICNARC: Intensive Care National Audit & Research Centre
NHS: National Health Service
UK: United Kingdom
PPE: Personal protective equipment
RR: Risk ratio
SARS-COV-2: Severe acute respiratory syndrome coronavirus 2
SOC: Standard Occupational Classification

## Declarations

### Ethics Approval

UK Biobank received ethical approval from the NHS National Research Ethics Service North West (11/NW/0382). All participants provided written informed consent before enrolment in the study, which was conducted in accordarce with the Declaration of Helsinki. The study protocol is available online (https://www.ukbiobank.ac.uk/wp-content/uploads/2011/11/UK-Biobank-Protocol.pdf).

### Consent for publication

Not applicable

### Availability of data and materials

The data that support the findings of this study are available from UK Biobank (https://www.ukbiobank.ac.uk/) but restrictions apply to their availability. These data were used under licence for the current study, and so are not publicly available. The data are available from the authors upon reasonable request and with permission of UK Biobank.

### Authors’ contributions

SVK, KOD and JPP conceived the idea for the paper. CLN conducted the analysis. All authors contributed to the interpretation of the findings. CLN and SVK jointly wrote the first draft. All authors critically revised the paper for intellectual content and approved the final version of the manuscript. The corresponding authors (SVK and CLN) had full access to all the data in the study and had final responsibility for the decision to submit for publication.

## Acknowledgements

We are grateful to UK Biobank participants. This research has been conducted using the UK Biobank resource under Application 41686.

## References

1. World Health Organization. Coronavirus disease 2019 (COVID-19): Situation Report – 91. Geneva, 2020.

2. Sattar N, McInnes IB, Jjv M. Obesity a risk factor for severe COVID-19 infection: multiple potential mechanisms. Circulation 2020: In press.

3. Zhou F, Yu T, Du R, et al. Clinical course and risk factors for mortality of adult inpatients with COVID-19 in Wuhan, China: a retrospective cohort study. The Lancet 2020; 395(10229): 1054–62.

4. Wu C, Chen X, Cai Y, et al. Risk Factors Associated With Acute Respiratory Distress Syndrome and Death in Patients With Coronavirus Disease 2019 Pneumonia in Wuhan, China. JAMA Internal Medicine 2020.

5. Myers EM. Compounding Health Risks and Increased Vulnerability to SARS-CoV-2 for Racial and Ethnic Minorities and Low Socioeconomic Status Individuals in the United States. Preprints 2020: 2020040234.

6. Hutchins SS, Fiscella K, Levine RS, Ompad DC, McDonald M. Protection of racial/ethnic minority populations during an influenza pandemic. Am J Public Health 2009; 99(S2): S261–S70.

7. Khunti K, Singh AK, Pareek M, Hanif W. Is ethnicity linked to incidence or outcomes of covid-19? BMJ 2020; 369: m1548.

8. Platt L. Ethnicity and family: Relationships within and between ethnic groups: An analysis using the Labour Force Survey. 2009.

9. Sudlow C, Gallacher J, Allen N, et al. UK biobank: an open access resource for identifying the causes of a wide range of complex diseases of middle and old age. PLoS Med 2015; 12(3).

10. Jacob A, Justine R, Naomi A, et al. Dynamic linkage of COVID-19 test results between Public Health England’s Second Generation Surveillance System and UK Biobank; 2020.

11. Bhopal RS, Gruer L, Cezard G, et al. Mortality, ethnicity, and country of birth on a national scale, 2001–2013: A retrospective cohort (Scottish Health and Ethnicity Linkage Study). PLoS Med 2018; 15(3): e1002515.

12. Townsend P. Deprivation. J Soc Policy 1987; 16(2): 125–46.

13. Hagenaars SP, Gale CR, Deary IJ, Harris SE. Cognitive ability and physical health: a Mendelian randomization study. Scientific Reports 2017; 7(1): 2651.

14. Honkaniemi H, Juárez SP, Katikireddi SV, Rostila M. Psychological distress by age at migration and duration of residence in Sweden. Soc Sci Med 2020; 250: 112869.

15. Jani BD, Hanlon P, Nicholl BI, et al. Relationship between multimorbidity, demographic factors and mortality: findings from the UK Biobank cohort. BMC Medicine 2019; 17(1): 74.

16. Zou G. A modified poisson regression approach to prospective studies with binary data. Am J Epidemiol 2004; 159(7): 702–6.

17. Bhopal RS. Migration, Ethnicity, Race, and Health in Multicultural Societies. Oxford: Oxford University Press; 2014.

18. Munafò MR, Tilling K, Taylor AE, Evans DM, Davey Smith G. Collider scope: when selection bias can substantially influence observed associations. Int J Epidemiol 2017: dyx206–dyx.

19. Batty GD, Gale CR, Kivimäki M, Deary IJ, Bell S. Comparison of risk factor associations in UK Biobank against representative, general population based studies with conventional response rates: prospective cohort study and individual participant meta-analysis. BMJ 2020; 368: m131.

20. Zhao X, Zhang B, Li P, et al. Incidence, clinical characteristics and prognostic factor of patients with COVID-19: a systematic review and meta-analysis. medRxiv 2020.

21. Denaxas S, Hemingway H, Shallcross L, et al. Estimating excess 1-year mortality from COVID-19 according to underlying conditions and age in England: a rapid analysis using NHS health records in 3.8 million adults; 2020.

22. Kurian AK, Cardarelli KM. Racial and ethnic differences in cardiovascular disease risk factors: a systematic review. Ethn Dis 2007; 17(1): 143.

23. Intensive Care National Audit & Research Centre. ICNARC report on COVID-19 in critical care. London, 2020.

24. CDC. Cases of Coronavirus Disease (COVID-19) in the U.S. 2020. https://www.cdc.gov/coronavirus/2019-ncov/cases-updates/cases-in-us.html (accessed 20/4/2020.

25. Nayak A, Islam SJ, Mehta A, et al. Impact of Social Vulnerability on COVID-19 Incidence and Outcomes in the United States. medRxiv 2020: 2020.04.10.20060962.

26. Katikireddi SV, Leyland AH, McKee M, Ralston K, Stuckler D. Patterns of mortality by occupation in the United Kingdom, 1991-2011: A comparative analysis of linked census-mortality records over time and place. Lancet Public Health 2017; 2(11): e501–e12.

27. Aldridge RW, Story A, Hwang SW, et al. Morbidity and mortality in homeless individuals, prisoners, sex workers, and individuals with substance use disorders in high-income countries: a systematic review and meta-analysis. The Lancet 2018; 391(10117): 241–50.

28. Abubakar I, Aldridge RW, Devakumar D, et al. The UCL & Lancet Commission on Migration and Health: the health of a world on the move. The Lancet 2018; 392(10164): 2606–54.

29. Chin MH, Walters AE, Cook SC, Huang ES. Interventions to reduce racial and ethnic disparities in health care. Med Care Res Rev 2007; 64(5 suppl): 7S–28S.

30. Katikireddi SV, Cezard G, Bhopal RS, et al. Assessment of health care, hospital admissions, and mortality by ethnicity: population-based cohort study of health-system performance in Scotland. The Lancet Public Health 2018; 3(5): e226–e36.

31. Douglas M, Katikireddi SV, Taulbut M, McKee M, McCartney G. Mitigating the wider health effects of COVID-19 pandemic response. BMJ 2020: In press.

